# Parental Income Gradients in Adult Health: A National Cohort Study

**DOI:** 10.1101/2020.12.17.20248400

**Authors:** Miriam Evensen, Søren Toksvig Klitkou, Mette C Tollånes, Jonas Minet Kinge, Torkild Hovde Lyngstad, Stein Emil Vollset, Simon Øverland

**Author notes:** **Correspondence to:** Miriam Evensen, Address: Norwegian Institute of Public Health, PO Box 222-Skøyen, 0213 Oslo, Norway.

## Abstract

**INTRODUCTION:** Disparities in health by adult income are well documented, but we know less about the childhood origins of health inequalities. This study examined the association between parental income in childhood and health in adulthood.

**METHODS:** We used administrative data on seven complete Norwegian birth cohorts born 1967-1973 (N = 429, 886) to estimate the association between parental income from birth to age 18, obtained from tax records available from 1967, linked with administrative registries on health. Health measures, observed between ages 39 to 43, were taken from registry data on consultations at primary health care services and hospitalizations and out-patient specialist consultations registered in the National Patient Registry.

**RESULTS:** Low parental income during childhood was associated with a 10, 2 percentage-point higher risk of overall disorders between the 5% highest (66.8%, CI 66.2-67.3) and lowest (77.2%, CI 76.6-77.8) parental income vigintiles. Absolute differences were largest for disorders related to musculoskeletal pain, injuries, and depression (7-9 percentage-point difference). There were also differences for hypertension (8%, CI 7.9-8.5 versus 4%, CI 4.1-4.7) and diabetes (3.2%, CI 3.0-3.4 versus 1.4%, CI 1.2-1.6), but smaller differences in consultations related to respiratory disorders (20.9% CI 20.4-21.5 versus 19.7% CI 19.2-20.3). Stratified analyses by other parental characteristics (education and marital status) and own adult characteristics (education and income) still showed a parental income gradient.

**CONCLUSIONS:** Low parental income in childhood was typically associated with a two-to-threefold increase in somatic and psychological disorders measured in adulthood, even in a setting with universal health care. This indicates that access alone is not enough to break intergenerational patterns of socioeconomic differences in health.

**WHAT IS KNOWN:**

- It is well documented that higher incomes among adults is positively associated with health and longevity, however, less is known about how childhood parental income is related to adult health.

**WHAT THIS STUDY ADDS:**

- We used administrative data on seven full Norwegian birth cohorts with information on parental income covering the whole childhood and a broad set of adult health measures taken from primary and secondary health service consultations.
- We demonstrate substantial associations between childhood parental income and overall measures of adult health, as well as for a large number of specific diagnoses, within a societal context where access to high-quality health care is universal.
- These gradients in adult health by childhood parental income did not vary substantively by other childhood circumstances, such as health at birth, mother’s marital status, and parental education.
- Despite individuals’ own completed education and adult income being strongly related to adult health, we found remaining adult health gradients by childhood parental income within all subpopulations stratified by own level of adult socioeconomic attainment.

## INTRODUCTION

Individuals with low income tend to have worse health than those with higher incomes and trends over the last decades reveal increasing health inequalities in many countries. (1–6) Strong socio-economic gradients in health are also found in Scandinavia, (7–9) despite universal access to publicly financed health services, free education and strong welfare-state institutions. (10) Moreover, economic inequalities are also on the rise in Scandinavia (11)—a development that may impact health inequalities both within and across generations.

Prior research suggests that childhood factors are important factors for adult health; however, most studies have focused on mortality outcomes, with less knowledge about health inequalities prior in adult life. (12–14) The second strand of research has documented strong associations between socio-economic conditions in childhood and adult health. However, this strand of research relies on survey data with limited health measures, typically with small sample sizes and lack of reliable measures of parental socio-economic status over time. (15,16)

From a life-course perspective, parental income gradients in adult health may reflect differences between children from low and high-income families in factors such as health at birth, family instability and parental human capital. (17–19) Prior research has shown that low-income children obtain less education and lower wages as adults, which constitute important determinants of health. (20,21) Less is known about the extent to which upward mobility in education and the labour market of low-income childhood origins is related to attenuated gradients in adult health by parental income or whether such gradients persist regardless of improved adult attainments.

In this study, we (i)examine the association between parental income in childhood and a wide range of disorders measured in adulthood using data on Norwegian birth cohorts born in 1967-73, and (ii) evaluate factors associated with social gradients in health using information on their childhood context and socio-economic attainments in adulthood

## METHODS

### Data sources and study population

We used data from Norwegian administrative registries: the Population Registry, the National Registry for Personal Taxpayers, the National Education Database, the Medical Birth Registry (MFR), the Norwegian Control and Reimbursement Database (KUHR) and the National Patient Registry (NPR). Unique (de-identified) personal identifiers allow for linkage between all registers, and between children and parents. Annual income data on the parents was available from 1967 whereas health data was available from 2006-16 from KUHR and from 2008-16 from NPR. We restricted the study population to the full Norwegian birth cohorts born 1967-73, to measure adult health outcomes at ages 39-43. An overview of data sources are given in Appendix Figure A1.

### Measures

#### Parental income measure

Measures of parental income were constructed from annually reported pensionable labour market earnings, which includes both wages and income from self-employment. Parental income was calculated as the sum of the mothers’ and fathers’ combined annual earnings averaged over the child’s age interval 0-18, with annual earnings adjusted to the 2016 Norwegian Consumer Price Index level. Then, we ranked all individuals within their same birth cohort by their childhood parental income and divided them in vigintiles (i.e., 20 groups where each group represents 5% of all individuals).

#### Measures of health from data in primary and specialist health care

Norway’s health care system is organized into primary and specialist health care. Primary health care comprises services such as consultations with primary care physicians, while specialist health care includes somatic and psychiatric hospitals and drug treatment centres. Diagnostic information from primary care is registered in the Norwegian Control and Reimbursement Database (KUHR) according to the International Classification of Primary Care (ICPC-2).(22)

We report results for adult health based on several measures of primary health care consultations. First, we measured the share of individuals who had one or more primary care consultation over a 5-year period. Second, we selected disorders with a diagnostic code (excluding symptoms) and selected the 15 most frequent diagnoses. We included consultations related to fear of specific diseases as an indicator of health behaviour. The selected disorders were summarized into a single overall measure referred to as any disorder. For an overview of specific codes, see Appendix Table A1.

As a robustness check, we used data from the National Patient Registry (NPR) covering the entire population of patients in specialist health care, to classify somatic outpatient treatment, somatic inpatient treatment (hospitalizations), and psychiatric specialist treatment (inpatient and outpatient) based on ICD-10 codes.

#### Measures of childhood health, parental socio-demographics and adult attainment

We report analyses stratified by selected measures of individuals’ childhood circumstances and adult attainments. To capture health at birth, we used a measure of low birthweight (individuals who were below 2500 grams at birth). Parental education was measured when the child was 16 years, by selecting the highest educational attainment level of either the mother or the father and categorized into three different levels: (1) less than upper secondary education (up to 11 years of completed education); (2) full secondary (12 years); and (3) tertiary (13 or more years). Mother’s marital status measures whether the mother was married when the child was aged 16.

To capture the individual’s own educational attainment in adulthood, we measured the highest completed level of education at age 35 in four categories: (1) below secondary (9 years); (2) full secondary (12 years); (3) tertiary short (13-14 years); and (4) tertiary long (15 years or more). Adult earnings were measured as the mean of own earnings overages 30-36 and were divided into quartiles. Adult family status was measured by combining information about marriage (married/not married) and parental status (have children/no children) measured by the latest follow up (i.e., age 43). We differentiate between individuals who are or have been married with own children up to this age versus all others (i.e., no children and/or not married).

### Statistical analysis

The aim of the analysis was to assess the association between parental income and a broad range of adult health diagnoses. For each adult health outcome, we estimate the following linear regression model:

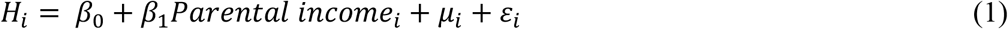

where *H*_*i*_ is the relevant health outcome of individual *i, parental income*_*i*_ is the measure of parental income rank and *μ*_*i*_is a set of birth cohort fixed effects. The parental income gradient in adult health is given by *β*_1_, which is the coefficient of parental income. To allow for non-linear gradients, we use a non-parametric specification where these gradients are from a set of dummy variables for parental income rank measured as vigintiles, and we report these results in a series of graphs.

All health outcomes are binary (i.e., 0 = no diagnosis, 1 = one or more consultations for with a diagnosis). We present the predicted probability of a given diagnosis by parental income vigintile from linear probability models (LPM) estimated using OLS regression. (23)

## RESULTS

Table 1 describes the sample. The study population consisted of 429,886 Norwegian individuals born in 1967-73. Descriptive statistics are shown separately for individuals from the lowest childhood parental income decile versus everyone else, which reveal that individuals from the lowest income origins have a higher prevalence for most adult health measures, they grew up with less-educated parents and in less stable families. As adults, they had lower income, less education, and were less likely to be married and have children.

**Table 1.** Descriptive statistics on adult health (age 39-43) and individual characteristics for Norwegian birth cohorts (1967-1973)

|  | All |  | Parental income,<br>decile 1 (lowest) |  | Parental income,<br>deciles 2-10 (highest) |  |
| --- | --- | --- | --- | --- | --- | --- |
|  | Mean | 95% CI | Mean | 95% CI | Mean | 95% CI, lower |
| **Measures from primary health care (KUHR)** |  |  |  |  |  |  |
| Any disorder | 0.739 | [0.737, 0.740] | 0.767 | [0.763, 0.771] | 0.735 | [0.734, 0.737] |
| Upper respiratory | 0.204 | [0.203, 0.206] | 0.208 | [0.204, 0.212] | 0.204 | [0.203, 0.205] |
| Acute sinusitis | 0.119 | [0.118, 0.120] | 0.117 | [0.114, 0.120] | 0.119 | [0.118, 0.120] |
| Back and neckpain | 0.168 | [0.167, 0.169] | 0.199 | [0.195, 0.203] | 0.165 | [0.164, 0.166] |
| Shoulder | 0.139 | [0.138, 0.140] | 0.157 | [0.154, 0.161] | 0.137 | [0.136, 0.138] |
| Bursitis | 0.116 | [0.115, 0.117] | 0.124 | [0.121, 0.127] | 0.115 | [0.114, 0.116] |
| Rheumatoid Arthritis (RA) | 0.013 | [0.013, 0.014] | 0.016 | [0.015, 0.017] | 0.013 | [0.013, 0.013] |
| Hypertension | 0.070 | [0.070, 0.071] | 0.081 | [0.078, 0.084] | 0.069 | [0.069, 0.070] |
| Overweight | 0.035 | [0.035, 0.036] | 0.042 | [0.040, 0.044] | 0.035 | [0.034, 0.035] |
| Diabetes | 0.024 | [0.023, 0.024] | 0.031 | [0.030, 0.033] | 0.023 | [0.022, 0.023] |
| Gastroenteritis infection | 0.010 | [0.009, 0.010] | 0.010 | [0.009, 0.011] | 0.009 | [0.009, 0.010] |
| Depression | 0.112 | [0.111, 0.113] | 0.142 | [0.138, 0.145] | 0.109 | [0.108, 0.109] |
| Anxiety | 0.040 | [0.040, 0.041] | 0.060 | [0.058, 0.063] | 0.038 | [0.038, 0.039] |
| Fear of specific diseases | 0.060 | [0.059, 0.061] | 0.064 | [0.062, 0.066] | 0.060 | [0.059, 0.060] |
| Naevus (mole) | 0.096 | [0.095, 0.097] | 0.083 | [0.080, 0.085] | 0.098 | [0.097, 0.099] |
| Injuries | 0.330 | [0.328, 0.331] | 0.359 | [0.354, 0.363] | 0.327 | [0.325, 0.328] |
| Any disorder (any of above) | 0.739 | [0.737, 0.740] | 0.767 | [0.763, 0.771] | 0.735 | [0.734, 0.737] |
| Any consultation in primary care | 0.955 | [0.954, 0.955] | 0.956 | [0.954, 0.958] | 0.954 | [0.953, 0.955] |
| **Measures from specialist health care (NPR)** |  |  |  |  |  |  |
| Outpatient | 0.466 | [0.465, 0.468] | 0.506 | [0.501, 0.510] | 0.462 | [0.460, 0.464] |
| Hospitalization, somatic | 0.170 | [0.169, 0.171] | 0.199 | [0.195, 0.203] | 0.167 | [0.165, 0.168] |
| Hospitalization, psychiatric | 0.075 | [0.074, 0.076] | 0.113 | [0.110, 0.116] | 0.071 | [0.070, 0.071] |
| **Individual measures from adulthood** |  |  |  |  |  |  |
| Adult income (age 30-36, NOK 2016) |  |  |  |  |  |  |
| Mean | 298,326 |  | 257,773 |  | 302,832 |  |
| Std. dev. | 364,183 |  | 115,977 |  | 381,663 |  |
| Education level (age 35) |  |  |  |  |  |  |
| Below secondary | 0.254 |  | 0.416 |  | 0.236 |  |
| Full secondary | 0.369 |  | 0.362 |  | 0.370 |  |
| University, short | 0.282 |  | 0.175 |  | 0.294 |  |
| University, long | 0.091 |  | 0.034 |  | 0.097 |  |
| Missing | 0.004 |  | 0.013 |  | 0.003 |  |
| Married with children | 0.461 |  | 0.370 |  | 0.472 |  |
| **Individual measures from childhood** |  |  |  |  |  |  |
| Parental income (age 0-18, NOK 2016) |  |  |  |  |  |  |
| Mean | 395,065 |  | 148,052 |  | 422,509 |  |
| Std. dev. | 151,890 |  | 64,537 |  | 132,811 |  |
| Parental education (age 16) |  |  |  |  |  |  |
| Less than full secondary | 0.567 |  | 0.839 |  | 0.537 |  |
| Full secondary | 0.195 |  | 0.098 |  | 0.256 |  |
| University | 0.237 |  | 0.057 |  | 0.257 |  |
| Missing | 0.001 |  | 0.057 |  | 0.006 |  |
| Mother not married at child age 16 | 0.154 |  | 0.365 |  | 0.131 |  |
| Mother's age at birth |  |  |  |  |  |  |
| Mean | 26.27 |  | 27.21 |  | 26.17 |  |
| Std. dev. | 5.54 |  | 7.09 |  | 5.33 |  |
| Low birth weight | 0.037 |  | 0.048 |  | 0.036 |  |
| First-born child of mother | 0.705 |  | 0.711 |  | 0.705 |  |
| Number of individuals | 429,886 |  | 42,987 |  | 386,899 |  |

### Childhood parental income, primary health care use, and number of diagnoses by chapter

Figure 1 (panel A) shows that the share of individuals with at least one consultation in primary health care is similar across parental income groups. Panel B gives an overview of the average number of consultations for ICPC-2 chapters of disorders across parental income groups. Individuals with low parental income generally had more consultations compared to individuals of higher-income origins and musculoskeletal and psychological disorders were the largest contributors to differences across income groups.

**Figure 1:**
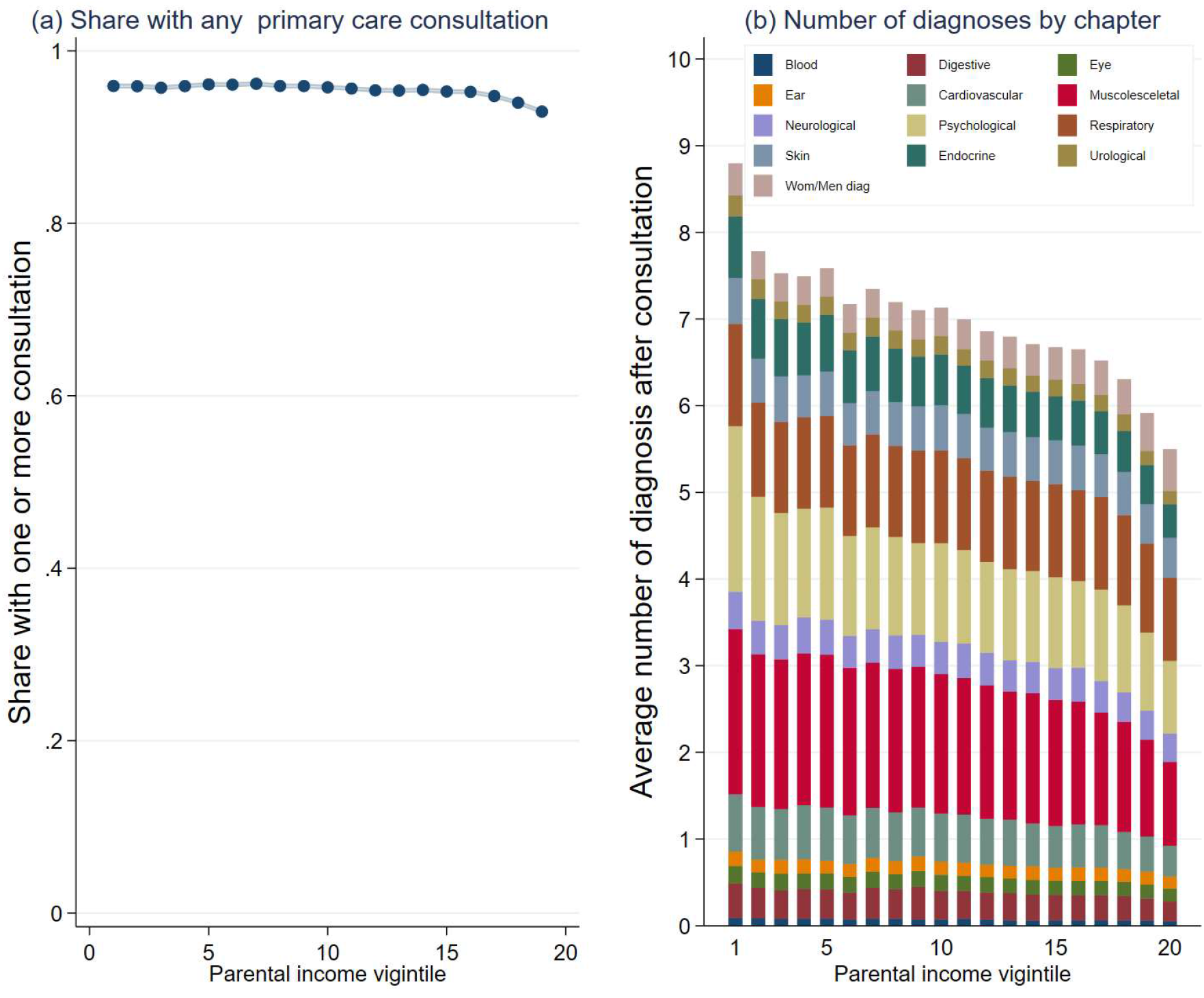
Share with any primary care consultation (panel A) and distribution of diagnosis chapters (panel B) by parental income vigintiles in childhood, Norwegian birth cohorts 1967-1975. *Source*: Data from the Norwegian Control and Reimbursement Database, 2006-2016. *Notes*: Childhood parental income vigintiles are averaged across the whole childhood, ages 0-18 years, and higher vigintiles refer to higher parental income. Panel A presents the share of individuals with one or more consultations in primary care in adulthood (ages 39-43) by childhood parental income vigintile. Panel B presents individuals’ average number of consultations in primary care for separate ICPC-2 chapters in adulthood (ages 39-43) by childhood parental income vigintile. We only include diagnoses within each category (excluding symptoms). Consultations related to Chapter A (General/unspecific) and W (Pregnancy-related disorders) are not included. Consultations from Chapters X (Female genital) and Y (Male genital) are combined (Wom/Men diag).

### Childhood parental income and cause-specific diagnoses in adulthood

Figure 2 shows the share with at least one diagnosed disorder (ages 39-43) by parental income vigintiles for the selected disorders (Table A2 report numerical values for point estimates and CIs). Higher parental income was associated with a lower share of disorders. For example, among individuals with parents in the lowest vigintile, 77.2% received a diagnosis (CI 76.6-77.8) compared to 66.8% among individuals in the highest vigintile (CI 66.2-67.3).

**Figure 2:**
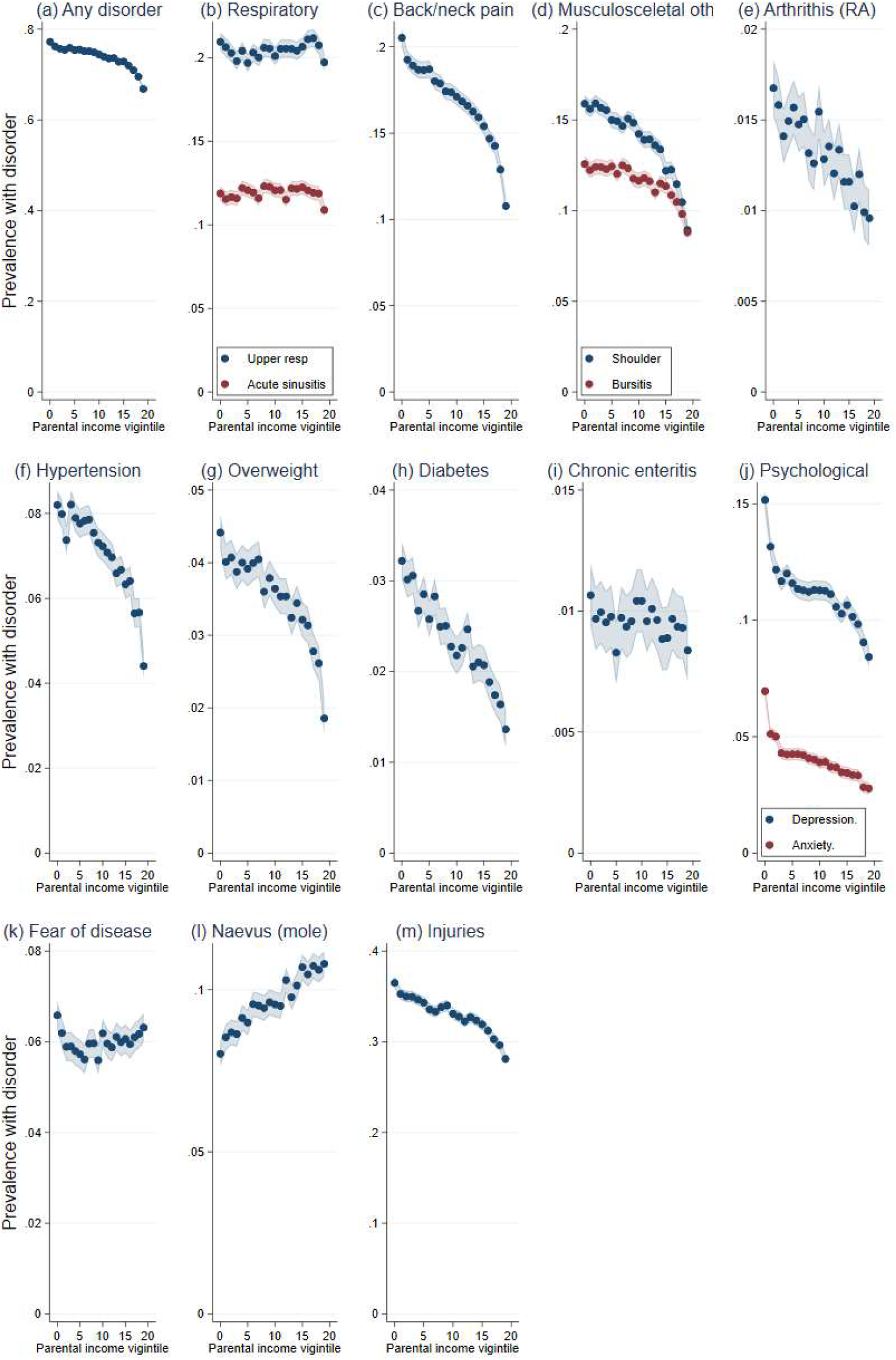
The association between parental income in childhood and adult health disorders in primary care, Norwegian birth cohorts 1967-1975. *Source*: Data from the Norwegian Control and Reimbursement Database, 2006-2016. *Note*: Predicted probabilities from linear probability models for childhood parental income vigintiles, controlling for birth year, estimated using OLS regression. Shaded areas refer to 95% confidence intervals. Panel A refers to the predicted probability of having received a diagnose for any of the specific disorders in panels B to M, while panels (B to M) refers to the predicted probabilities of having had one or more consultations for each of the specific disorders.

For specific diagnoses, there was no difference between income groups for respiratory disorders. The largest gap between individuals from the lowest and highest vigintile, in absolute terms, was found for back and neck pain and injuries. For example, there was a 9 percentage point higher probability of diagnosis with back and neck pain disorders among individuals with parents in the lowest income vigintile (20.5%, CI 20.0-21.0) compared to the highest vigintile group (10.8%, CI 10.3-11.3). For injuries, 36.5% (CI 35.9-37.1) among those in the lowest 5 % income group had an injury versus 28.1 % (CI 27.5-28.7) among the highest 5 %.

For chronic disorders, such as diabetes and overweight there were gradients, for the most part linear, where those in the top versus bottom vigintile generally had 2-3 percentage points higher probability of disorders. The overall prevalence of these disorders was low (in the range of 1-4 percent). For hypertension, the gap between the lowest (8.2%, CI 7.9-8.5) and highest (4.4%, CI 4.1-4.7) parental income vigintile was about 4 percentage points.

Parental income was strongly correlated with psychological disorders. For depression, there was a seven-percentage point difference between those in the lowest (15.2%, CI 14.8-15.6) versus highest vigintile (8.4%, CI 8.0-8.9). Anxiety disorders the differences between those in the lowest vigintile (7%, CI 6.7-7.2) compared to highest vigintile (2.8%, CI 2.5-3.0) was four percentage points.

For consultations related to mole and fear, were “fear of hypertension” and “fear of breast cancer” was among the most frequent, there was an inverse gradient, where higher parental income in childhood was associated with more consultations.

Women have more consultations than men, but the shape of the gradients are similar across sex (Appendix Figures A2, A3, and A4). The largest sex difference was found for injuries, with a weaker association across parental income for women than men (Appendix Figure A4).

### Sociodemographic variation in the gradients of adult health by childhood parental income

Figure 3 presents results for the summary measure of any disorders stratified by childhood circumstances (panels A-C) and adult attainments (panels D-F). We found a higher prevalence of disorders among individuals with low birth weight, although the differences were small and sometimes overlapping (panel A). There was also a higher prevalence of disorders among children in single-parent households relative to those with married parents, but the shape of the gradient was similar (panel B). Children of less-educated parents had a higher prevalence of disorders compared to those who had parents with tertiary university-level education (panel C).

**Figure 3:**
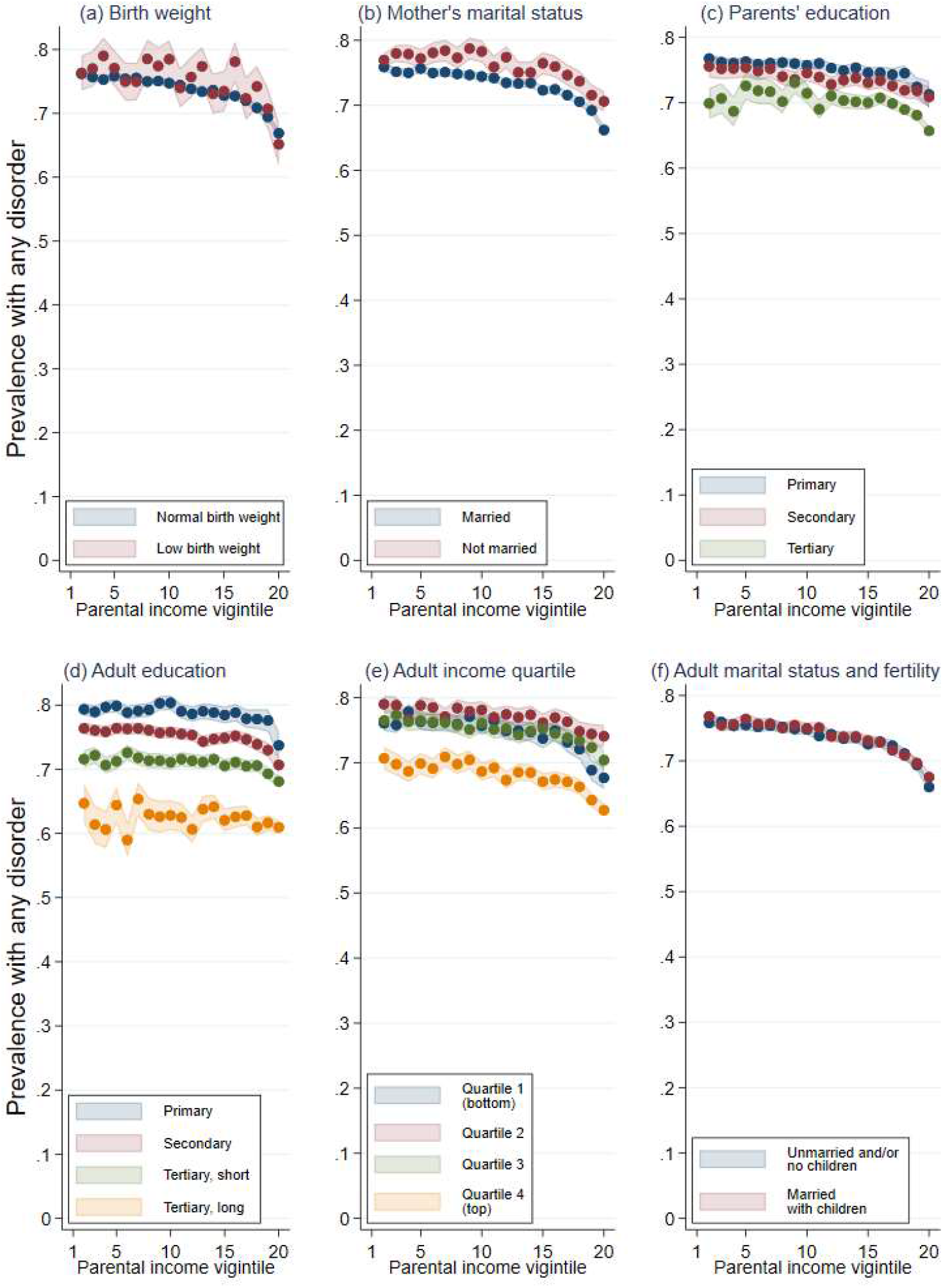
The association between parental income across childhood and adult health separately by childhood circumstances (panels A to C) and adult socio-economic attainment (panels D to F), Norwegian birth cohorts 1967-1975. *Source*: Data from the Norwegian Control and Reimbursement Database, 2006-2016. *Note*: Predicted probabilities of having had one or more consultations for any disorder (i.e., the same as in panel A in Figure 2) from linear probability models for childhood parental income vigintiles, controlling for birth year, estimated using OLS regression. Shaded areas refer to 95% confidence intervals. Each panel shows the predicted probabilities from regressions where each childhood parental income vigintile is separately interacted with dummy variable measures of birth weight (panel A), mother’s marital status at child age 16 (panel B), parental education at child age 16 (panel C), educational attainment at age 35 (panel D), adult income quartile averaged across ages 30-36 (panel E), and has been or being married with own children by age 43 (panel F).

Turning to adult attainment, we find marked differences in the prevalence of any disorder by their educational attainment (panel D). For example, for individuals who grew up in families in the lowest vigintile of the parental income distribution, the difference between those who themselves had completed less than upper secondary education or long tertiary education is 23.5 percentage points (p<0.001). Nonetheless, there is a childhood parental income gradient within all educational levels, although this gradient is less stable among those with long tertiary education. There are also parental income gradients in adult health within all quartiles of their adult income (panel E). However, those in the highest adult income quartile have better health than those found in other parts of the income distribution. Finally, there are small differences in health between those who are both married and have children of their own compared to all others (panel F).

### Robustness check

One may raise the concern that consultations in primary health care do not measure the most severe disorders and health conditions, although access to the more selective consultations in the specialist health care usually goes through the referral of a general practitioner. To provide a robustness check of the results for primary health care service use, we report results for somatic outpatient consultations, somatic inpatient consultations (hospitalizations) and psychiatric consultations (outpatient and inpatient consultations) in the specialist health care using data from the National Patient Registry.

Figure 4 reveals a similar pattern as reported for primary health care above, showing a relatively linear gradient where lower parental income is associated with higher probabilities of both outpatient and inpatient treatments of somatic conditions. For psychiatric treatments, the association strongly resembles the shape found in Figure 2 (panel J) for anxiety and depression with a more pronounced tail, with more than a twofold increased risk between individuals from the highest and lowest childhood income origins.

**Figure 4:**
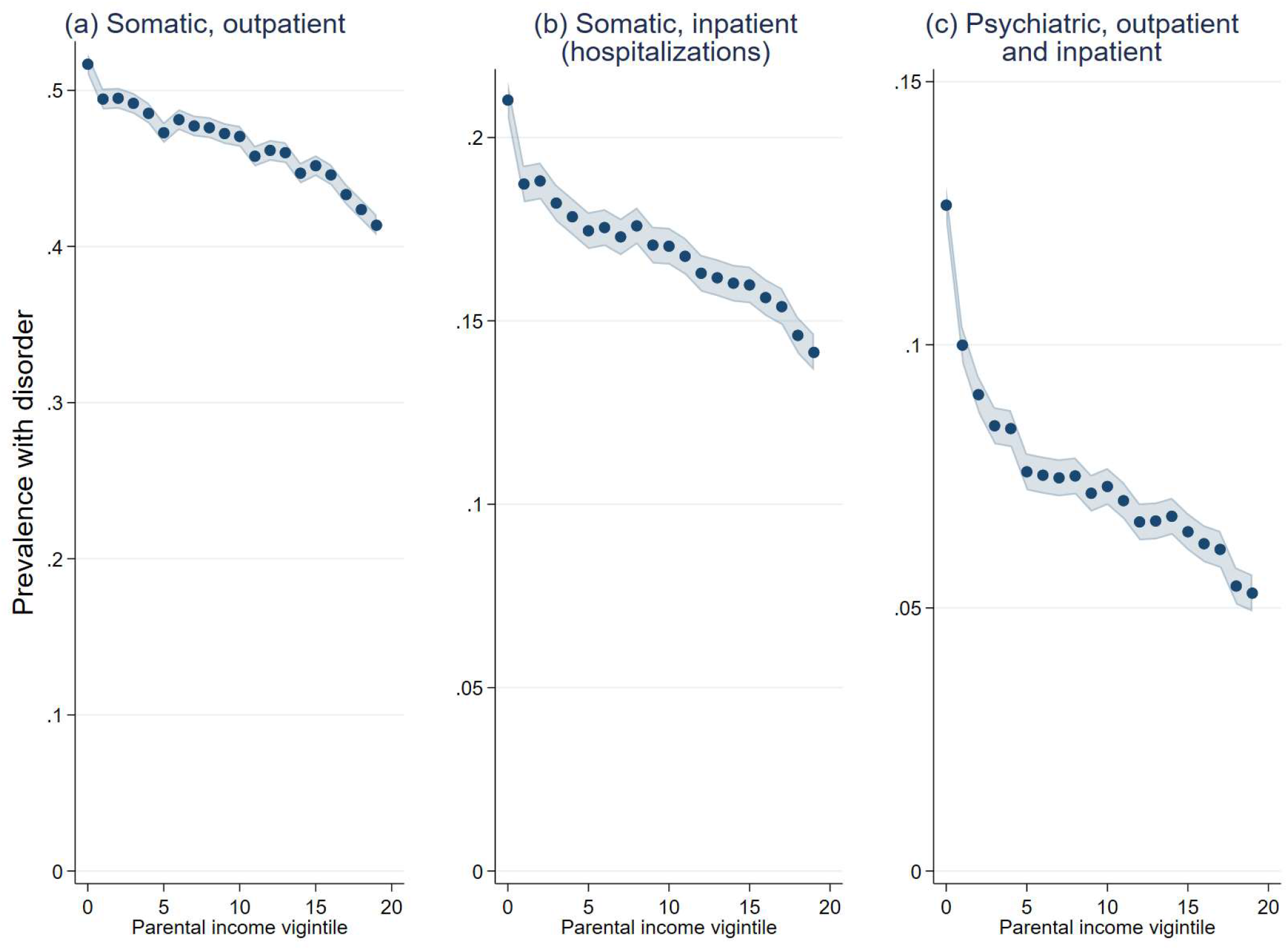
The association between parental income and health outcomes in specialist health care, Norwegian birth cohorts 1967-1975. *Source*: Data from the National Patient Registry, 2008-2016. *Note*: Predicted probabilities from linear probability models for childhood parental income vigintiles, controlling for birth year, estimated using OLS regression. Shaded areas refer to 95% confidence intervals. Panel A shows the predicted probability of having had one or more somatic outpatient consultations in the specialist health care between ages 41-43 years. Panel B shows the predicted probability of having had one or more somatic inpatient (hospitalization) consultations in the specialist health care between ages 41-43 years. Panel C shows the predicted probability of having had one or more psychiatric consultations (including both outpatient and inpatient) in the specialist health care between ages 41-43 years.

## DISCUSSION

In this study, we used population-wide data to obtain estimates of social gradients in health for a broad range of disorders as registered in primary and specialist health care. Further, we identified factors related to family resources and childhood upbringing that were correlated with social gradients in health. We found that parental income in childhood is related to adult health status for a wide range of disorders. The largest absolute difference was observed for disorders related to injuries, psychological and musculoskeletal disorders with a doubling of the risk of these disorders between individuals who grew up in the bottom and top vigintile of the parental income distribution. For diabetes and overweight, although less common in these age groups, there was a two-to threefold increase in disorders.

The documented health gradients are substantial and particularly so given that the study population was relatively young. Not only are these individuals expected to reach older age without their health intact, but these health differences may also have consequences for work capacity and retirement decisions. (24,25) Our results corroborate smaller studies documenting a higher risk of disorders among individual from low-income backgrounds that primarily rely on self-reported measures (26), but we examine a broader range of disorders using administrative data. A higher share of injuries among those from low-income families is also consistent with previous findings showing higher mortality rates due to accidents for this group. (27)

These differences are unlikely tied to access to medical care as we found smaller differences in overall share of primary care consultations between individuals from high and low-income origins (28). However, even though access to care is universal, the quality of care may differ by socio-economic background. For consultations related to mole and fear of specific diseases, we found an inverse relationship where higher childhood parental income was related to more consultations. This might indicate that individuals from high-income origins engage in more preventive and risk control behaviour consistent with prior research showing socio-economic gradients in, e.g., mammogram participation and medication adherence. (29,30)

Children growing up in low-income families face disadvantages along several additional dimensions, such as prenatal health, family instability, and parents’ lack of educational resources (19,31–33). One of our key findings is that adult health gradients by childhood parental income are relatively similar within different subpopulations stratified by differences in childhood circumstances. Parental educational attainment was strongly related to adult health, indicating that low income in childhood is more severe if parents lack additional resources to compensate for lack of income.

By contrast, own adult attainments may attenuate the link between individuals’ adult health and childhood income origins. In this regard, the pattern observed for adult educational attainment was particularly revealing. For those from low-income family backgrounds, who – against odds – attained a high level of education, we still found a similar shape of the gradient. This indicates that those who are upwardly mobile in terms of educational attainment are not able to fully overcome the disadvantages of low-income childhood origins. For adult income, we found a parental income gradient within all quartiles of own adult income, but the difference between the highest and lowest quartile in own income seems to be of the same magnitude as the overall gradient by childhood parental income. Differences in own family formation were less related to differences in adult health. However, problems of reverse causality might be important for all adult attainments, as individuals might fare less well because of health problems rather than the reverse.

There are strengths and limitations with the approach used here. Unlike most prior research, which uses data from self-reports or hospitalizations or mortality, we rely on physician-registered health disorders, a measure that might complement other measures of health. However, the validity of accurate diagnoses may be questioned as research has shown that physicians differ in their evaluation of health problems. (34,35) For the broader groups of disorders related to psychological and musculoskeletal problems with uncertain symptoms, differences in physicians’ assessment may influence the results. Still, if there are systematic differences in the assessment of symptoms, our use of multiple health disorders might mitigate this bias. Our results are also limited to individuals who have been in contact with primary or secondary care services. If the tendency to seek medical help varies by parental income, this may bias the association.

Another caveat is related to the age of the sample. By focusing on health at age 39-43, we likely capture some of the chronic diseases that begin to emerge and impair functional abilities, but we may also miss disorders with later onsets. For example, we found no relationship to respiratory disorders, which is surprising given the relation of income to smoking and physical activity. (6) However, smoking rates have drastically declined across generations implying that such factors are less relevant for these and younger cohorts. (36) Instead, other worrisome trends have emerged, such as increases in childhood obesity (37) as well as high rates of mental disorders among children and adolescents. (38,39) How these trends will affect health disparities among the coming generations are a topic for future research.

To conclude, we found substantial associations between childhood parental income and adult health, particularly for injuries, psychological, and musculoskeletal disorders. These disorders often have a recurring trajectory and have implications for years lived in good health. Access to free health care, although important for many reasons, are not enough to overcome these differences. These results can inform policy development intending to advance health equity.

## Data Availability

Data are not publicly available; however, they can be accessed by Statistics Norway and relevant government agencies upon relevant approvals.

