## Supplemental materials for "Parental Income Gradients in Adult Health: A National Cohort Study"

Supplementary Materials for Parental Income Gradients in Adult Health: A National Cohort Study.

This file includes details on coding, construction of demographic and socioeconomic variables and sensitivity analyses and robustness checks, Tables A1-A3, Figure A1-A6.

**<u>Supplementary tables:</u>**

Table A1: Codes for categorizing disorders
Table A2: Numerical values for Figure 2, 95% CI and coefficients
Table A3: Comparison of mothers union status by birth and at child age 16

**<u>Supplementary Figures:</u>**

Figure A1: Study design and data sources, Norwegian birth cohorts 1967-1975
Figure A2: Number of consultations across 5-years (panel A) and share with any primary care consultation (panel B) by parental income percentiles in childhood, Norwegian birth cohorts 1967-1975.
Figure A3: The association between parental income in childhood and diagnosed disorders by each ICPC-2 chapter in primary care, Norwegian birth cohorts 1967-1975.
Figure A4: The association between parental income in childhood and adult health disorders in primary care, Norwegian birth cohorts 1967-1975.
Figure A5: Share with any disorder for separate measures of mother and father income rank in childhood (panel A) and share with any disorder separate by mother's marital status (panel B)
Figure A6: Unadjusted and adjusted association between parental income in percentiles in childhood and adult health (any disorder) at age 39-43, Norwegian birth cohorts 1967-1975

Figure A1: Study design and data sources, Norwegian birth cohorts 1967-1975

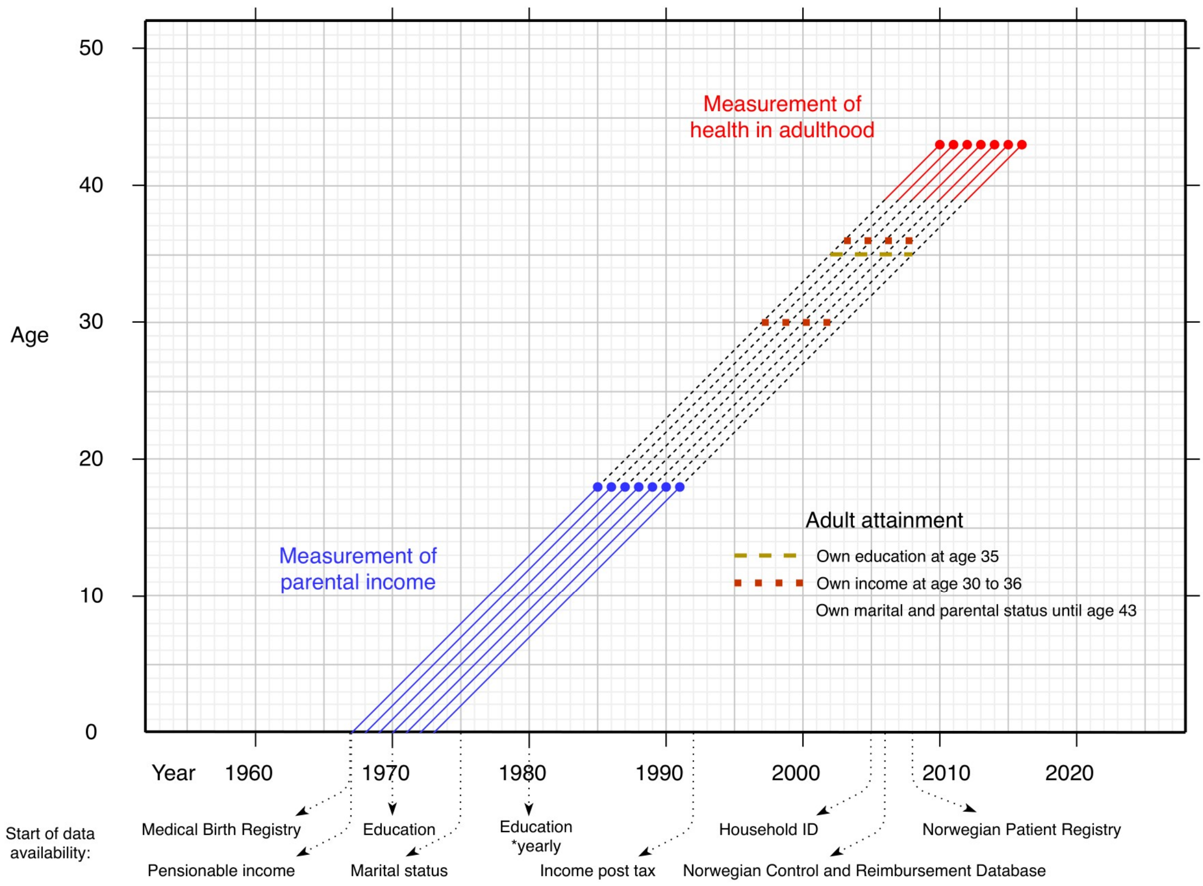


**Details on coding and reporting of health variables**

To measure health in this study, individual level data on diagnoses were taken from different sources. All residents in Norway are members of the National Insurance Scheme and are assigned a general practioner. Primary care providers (mainly physician and emergency care units and contracting specialists care) report reimbursement code which are recorded in the Norwegian Control and Reimbursement Database (KUHR), available from 2006 and onwards, Diagnostic information is mandatory to report for each patient contact and are unlikely to go unreported since these reporting are essential for reimbursement. Primary care services are coded according to the *International Classification of Primary Care, second edition* (ICPC-2). Some of the registrations in KUHR does not necessarily reflect an in- person contact (they may be advice given by letter/phone/ prescriptions/electronic contact etc.) However, to measure any consultation or disorder, we rely on information from personal consultations identified based on tariff rates.

The Norwegian Patient Registry (NPR) is a nationwide health register covering all out- and inpatient specialist services in Norway, available from 2008 and onwards. The diagnoses are coded according to the International Classification of diseases and related health problem, 10th edition. Usually, a reference from the general practioner is necessary to get specialist treatment, (except for acute hospitalization and treatment at emergency department). While the majority of injuries are reported in primary care, some emergency care units, report injuries exclusively to NPR and thus are coded according to the ICD-10 classification. (1) Table A1 provide an overview of the specific codes used from ICPC-2 and ICD-10.

Table A1: Code for categorizing disorders from ICPC-2 and ICD-10

| | ICPC-2 | ICD-10 |
|---|---|---|
| Back and neckpain | L83, L84, L86 | |
| Shoulder syndrome | L92, L93 | |
| Bursitis | L87 | |
| Depression | P76 | |
| Anxiety | P74 | |
| Hypertension | K86-K87 | |
| Diabetes | T90 | |
| Overweight | T84 | |
| Artrithis | L88 | |
| Fear of specific diseases | A25-A27, B25-B27, D26-D27, F27, H27, K24-K27, L26-L27, N26-N27, P27, R26-R27, S26-S27, T26-T27, U26-U27, X23-X27, Y24-Y27 | |
| Chronic enteritis | D94 | |
| Naevous | S82 | |
| Injuries and accidents | A80-A82, A84, A86, A88, B76-B77, D79-D80, F75-F79, H76-H79, L72-L81, L96, N79-N81, R87-R88, S12-S19, S80, U80, X82, Y80 | S00-T78 and V01-Y36 |

Note: While most emergency units report injuries to the the national registry for reimbursemnet of primary care providers (KUHR) some emergency care units report injuries to the National Patient Registry. We therefore included codes from ICPC-2 and ICD-10. First listed diagnosis was used.

**Figure A2:** Number of consultations across 5-years (panel A) and share with any primary care consultation (panel B) by parental income percentiles in childhood, Norwegian birth cohorts 1967-1975.

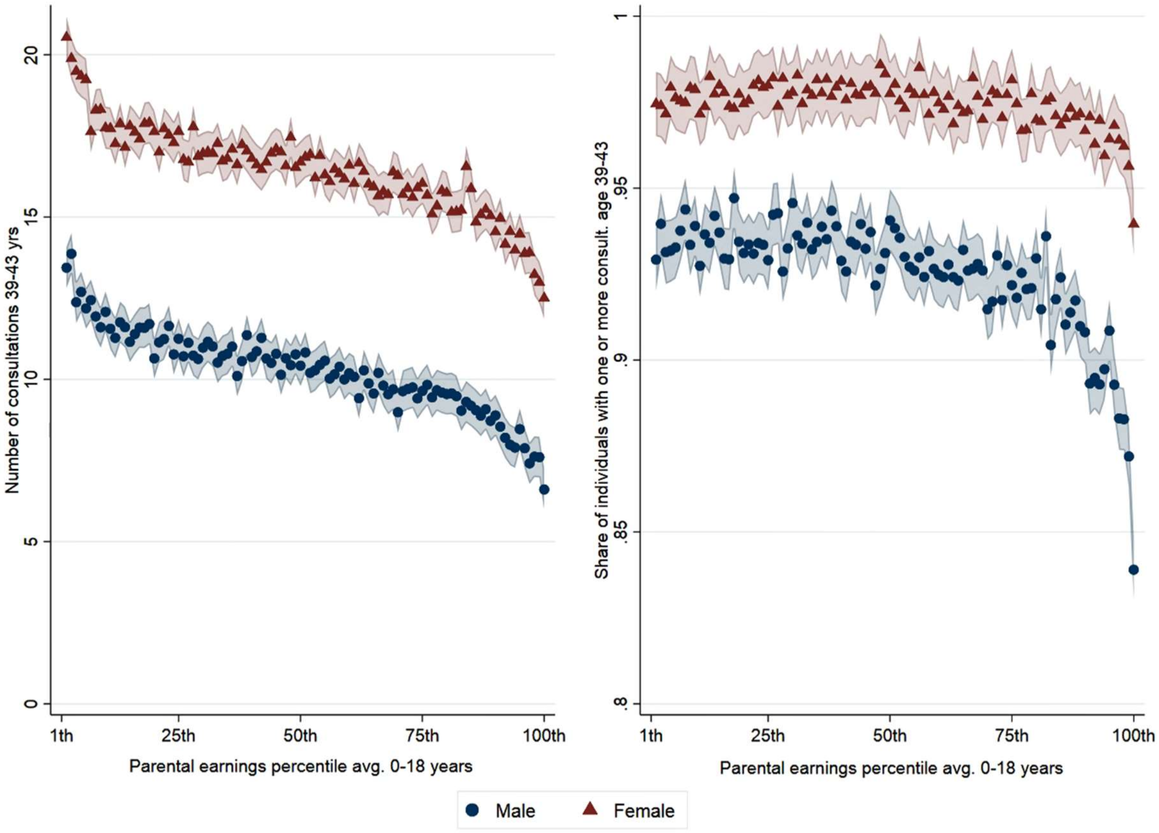


*Source*: Data from the Norwegian Control and Reimbursement Database, 2006-2016.

*Notes*: Childhood parental income percentiles are averaged across the whole childhood, ages 0-18 years, and higher percentiles refer to higher parental income. Panel A presents the average number of consultations in primary care in adulthood (ages 39-43). Panel B presents the share of individuals with one or more consultations in primary care in adulthood (ages 39-43) by childhood parental income percentile.

**Figure A3:** The association between parental income in childhood and diagnosed disorders by each ICPC-2 chapter in primary care, Norwegian birth cohorts 1967-1975.

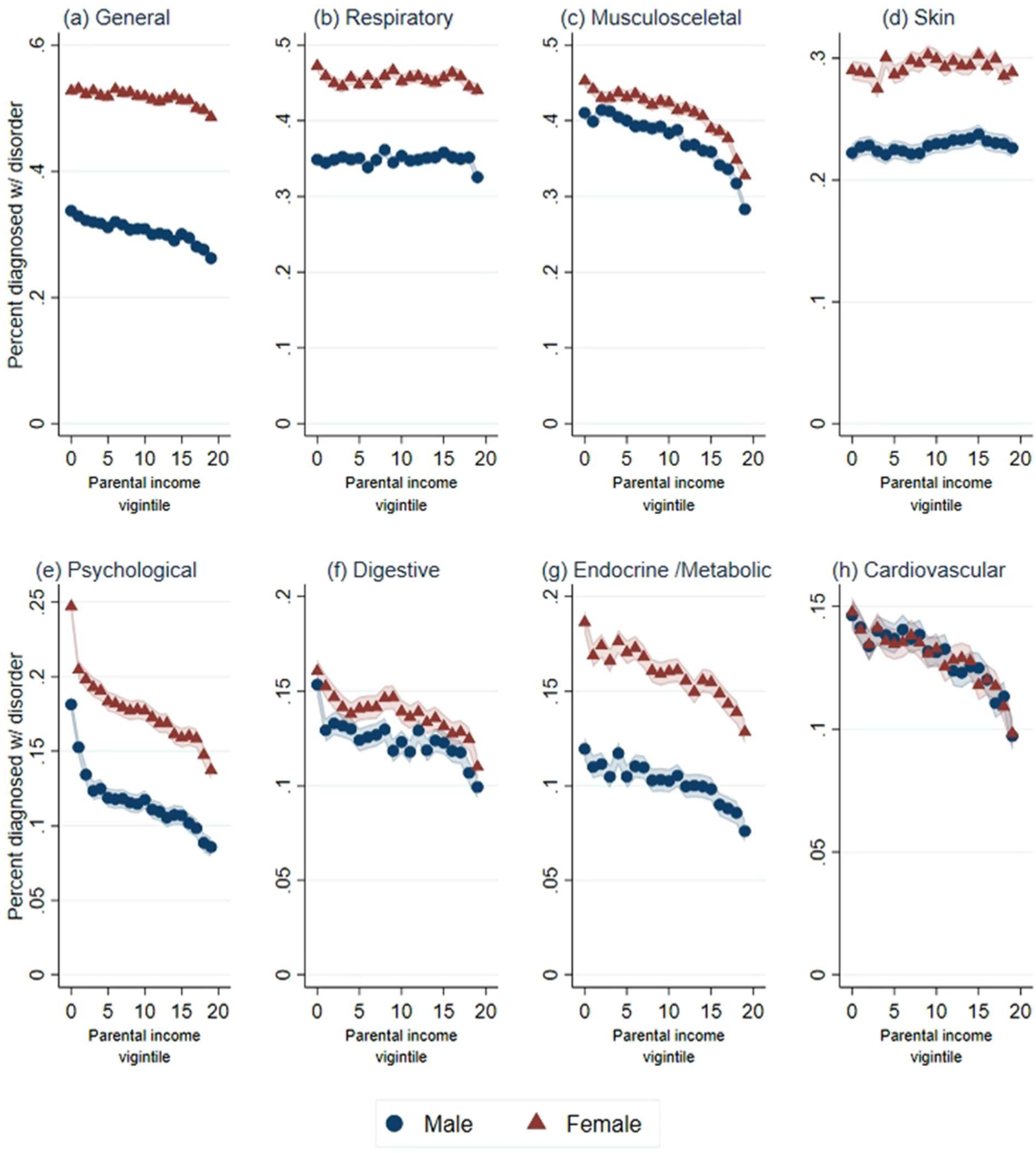


(continues on next page)

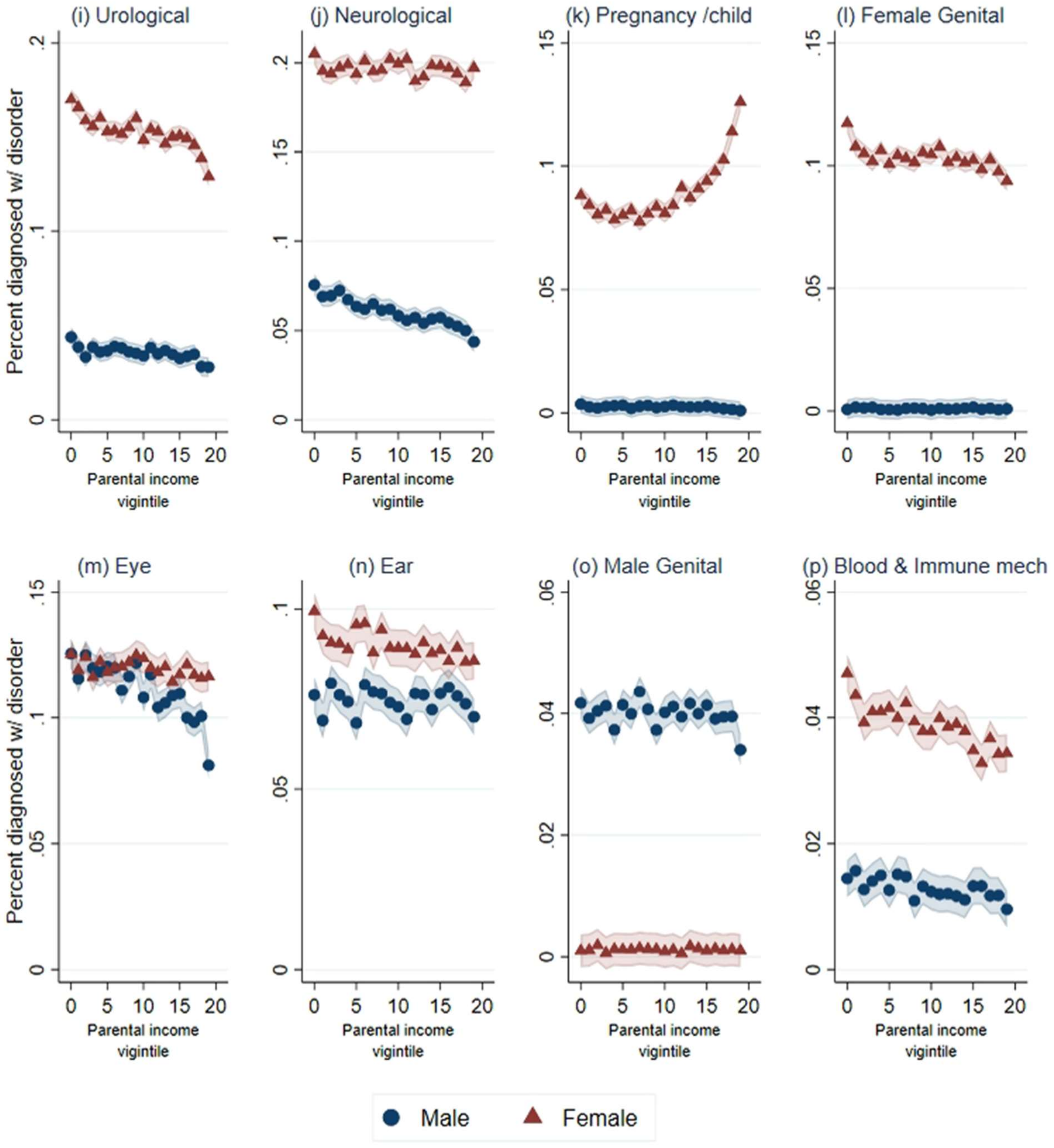


*Source*: Data from the Norwegian Control and Reimbursement Database, 2006-2016.

*Note*: Predicted probabilities from linear probability models for childhood parental income vigintiles, controlling for birth year, estimated using OLS regression. Shaded areas refer to 95% confidence intervals.

Figure A4: The association between parental income in childhood and adult health disorders in primary care, Norwegian birth cohorts 1967-1975.

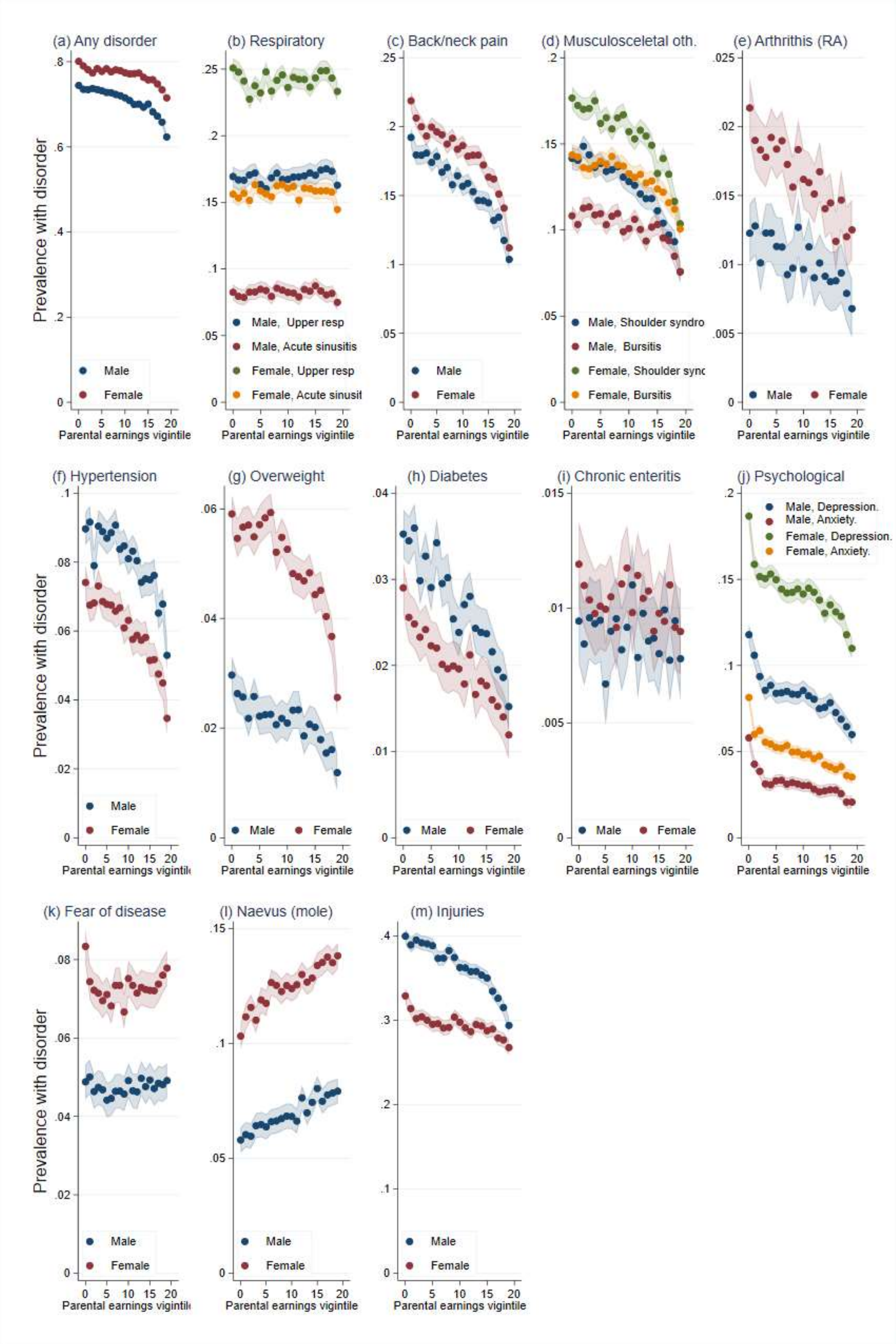


*Source*: Data from the Norwegian Control and Reimbursement Database, 2006-2016.

*Note*: Predicted probabilities from linear probability models for childhood parental income vigintiles, controlling for birth year, estimated using OLS regression. Shaded areas refer to 95% confidence intervals.

Table A2: Nummerical values and 95% CI for Figure 2

| | Any disorder | | Upper respiratory | | Sinusitis | | Back and neck pain | | Shoulder pain | | Bursitis | | Arthrithis (RA) | | Overweight | |
|---|---|---|---|---|---|---|---|---|---|---|---|---|---|---|---|---|
| Vigintiles | Prev. | 95% CI | Prev. | 95% CI | Prev. | 95% CI | Prev. | 95% CI | Prev. | 95% CI | Prev. | 95% CI | Prev. | 95% CI | Prev. | 95% CI |
| 1st (lowest) | 0.772 | [0.766,0.778] | 0.209 | [0.204,0.215] | 0.119 | [0.114,0.123] | 0.205 | [0.200,0.210] | 0.159 | [0.154,0.163] | 0.126 | [0.121,0.130] | 0.017 | [0.015,0.018] | 0.044 | [0.042,0.047] |
| 2 | 0.762 | [0.756,0.768] | 0.206 | [0.201,0.212] | 0.115 | [0.111,0.120] | 0.193 | [0.188,0.198] | 0.156 | [0.151,0.161] | 0.122 | [0.118,0.127] | 0.016 | [0.014,0.017] | 0.040 | [0.038,0.043] |
| 3 | 0.757 | [0.752,0.763] | 0.203 | [0.197,0.208] | 0.117 | [0.112,0.121] | 0.189 | [0.184,0.194] | 0.159 | [0.154,0.164] | 0.124 | [0.120,0.128] | 0.014 | [0.013,0.016] | 0.041 | [0.038,0.043] |
| 4 | 0.755 | [0.749,0.760] | 0.198 | [0.193,0.203] | 0.116 | [0.111,0.120] | 0.187 | [0.182,0.192] | 0.157 | [0.152,0.161] | 0.124 | [0.120,0.128] | 0.015 | [0.013,0.016] | 0.039 | [0.036,0.041] |
| 5 | 0.759 | [0.753,0.765] | 0.204 | [0.199,0.209] | 0.122 | [0.118,0.126] | 0.187 | [0.182,0.192] | 0.155 | [0.151,0.160] | 0.123 | [0.119,0.127] | 0.016 | [0.014,0.017] | 0.040 | [0.038,0.042] |
| 6 | 0.754 | [0.748,0.760] | 0.197 | [0.191,0.202] | 0.121 | [0.116,0.125] | 0.187 | [0.182,0.192] | 0.150 | [0.145,0.155] | 0.124 | [0.120,0.129] | 0.015 | [0.013,0.016] | 0.039 | [0.037,0.042] |
| 7 | 0.755 | [0.749,0.761] | 0.203 | [0.198,0.208] | 0.119 | [0.115,0.124] | 0.180 | [0.175,0.185] | 0.149 | [0.145,0.154] | 0.120 | [0.116,0.125] | 0.015 | [0.013,0.017] | 0.040 | [0.037,0.042] |
| 8 | 0.751 | [0.745,0.757] | 0.200 | [0.195,0.206] | 0.116 | [0.112,0.120] | 0.179 | [0.174,0.184] | 0.147 | [0.142,0.151] | 0.125 | [0.121,0.129] | 0.013 | [0.012,0.015] | 0.040 | [0.038,0.043] |
| 9 | 0.751 | [0.746,0.757] | 0.206 | [0.201,0.211] | 0.123 | [0.119,0.127] | 0.174 | [0.169,0.179] | 0.151 | [0.146,0.155] | 0.123 | [0.119,0.128] | 0.013 | [0.011,0.014] | 0.036 | [0.034,0.038] |
| 10 | 0.749 | [0.743,0.755] | 0.205 | [0.200,0.211] | 0.123 | [0.118,0.127] | 0.174 | [0.169,0.179] | 0.148 | [0.144,0.153] | 0.118 | [0.113,0.122] | 0.015 | [0.014,0.017] | 0.038 | [0.035,0.040] |
| 11 | 0.744 | [0.738,0.750] | 0.201 | [0.196,0.206] | 0.121 | [0.116,0.125] | 0.171 | [0.166,0.176] | 0.142 | [0.138,0.147] | 0.116 | [0.112,0.121] | 0.013 | [0.011,0.014] | 0.036 | [0.034,0.039] |
| 12 | 0.739 | [0.733,0.745] | 0.205 | [0.200,0.211] | 0.121 | [0.116,0.125] | 0.168 | [0.163,0.173] | 0.139 | [0.134,0.144] | 0.118 | [0.114,0.122] | 0.014 | [0.012,0.015] | 0.035 | [0.033,0.038] |
| 13 | 0.735 | [0.730,0.741] | 0.205 | [0.200,0.211] | 0.115 | [0.111,0.119] | 0.166 | [0.161,0.171] | 0.139 | [0.135,0.144] | 0.116 | [0.112,0.120] | 0.012 | [0.011,0.014] | 0.035 | [0.033,0.038] |
| 14 | 0.736 | [0.730,0.742] | 0.205 | [0.200,0.211] | 0.122 | [0.118,0.126] | 0.163 | [0.158,0.168] | 0.136 | [0.131,0.141] | 0.110 | [0.106,0.114] | 0.013 | [0.012,0.015] | 0.032 | [0.030,0.035] |
| 15 | 0.728 | [0.722,0.734] | 0.204 | [0.199,0.209] | 0.122 | [0.117,0.126] | 0.159 | [0.154,0.164] | 0.134 | [0.129,0.138] | 0.115 | [0.111,0.119] | 0.012 | [0.010,0.013] | 0.034 | [0.032,0.037] |
| 16 | 0.729 | [0.723,0.735] | 0.207 | [0.201,0.212] | 0.123 | [0.118,0.127] | 0.154 | [0.149,0.159] | 0.122 | [0.117,0.127] | 0.113 | [0.109,0.118] | 0.012 | [0.010,0.013] | 0.032 | [0.030,0.035] |
| 17 | 0.720 | [0.714,0.726] | 0.211 | [0.206,0.216] | 0.121 | [0.116,0.125] | 0.147 | [0.142,0.152] | 0.123 | [0.118,0.127] | 0.109 | [0.104,0.113] | 0.010 | [0.009,0.012] | 0.031 | [0.029,0.034] |
| 18 | 0.710 | [0.704,0.716] | 0.212 | [0.206,0.217] | 0.119 | [0.115,0.124] | 0.143 | [0.138,0.148] | 0.115 | [0.110,0.119] | 0.105 | [0.100,0.109] | 0.012 | [0.010,0.014] | 0.028 | [0.025,0.030] |
| 19 | 0.695 | [0.689,0.701] | 0.207 | [0.202,0.213] | 0.119 | [0.114,0.123] | 0.129 | [0.124,0.134] | 0.105 | [0.100,0.109] | 0.098 | [0.094,0.102] | 0.010 | [0.008,0.011] | 0.026 | [0.024,0.029] |
| 20th (highest) | 0.668 | [0.662,0.674] | 0.197 | [0.192,0.203] | 0.109 | [0.105,0.113] | 0.108 | [0.103,0.113] | 0.089 | [0.085,0.094] | 0.088 | [0.084,0.092] | 0.010 | [0.008,0.011] | 0.019 | [0.016,0.021] |

| | Hypertension | | Diabetes | | Depression | | Anxiety | | Fear of disorders | | Naevus/mole | | Enteritis | | Injuries | |
|---|---|---|---|---|---|---|---|---|---|---|---|---|---|---|---|---|
| | Prev. | 95% CI | Prev. | 95% CI | Prev. | 95% CI | Prev. | 95% CI | Prev. | 95% CI | Prev. | 95% CI | Prev. | 95% CI | Prev. | 95% CI |
| 1st (lowest) | 0.082 | [0.079,0.085] | 0.032 | [0.030,0.034] | 0.152 | [0.148,0.156] | 0.070 | [0.067,0.072] | 0.066 | [0.063,0.069] | 0.080 | [0.076,0.084] | 0.011 | [0.009,0.012] | 0.365 | [0.359,0.371] |
| 2 | 0.080 | [0.076,0.083] | 0.030 | [0.028,0.032] | 0.132 | [0.127,0.136] | 0.051 | [0.049,0.054] | 0.062 | [0.059,0.065] | 0.085 | [0.081,0.089] | 0.010 | [0.008,0.011] | 0.353 | [0.346,0.359] |
| 3 | 0.074 | [0.070,0.077] | 0.031 | [0.029,0.033] | 0.122 | [0.118,0.126] | 0.050 | [0.047,0.053] | 0.059 | [0.056,0.062] | 0.087 | [0.083,0.091] | 0.010 | [0.009,0.011] | 0.350 | [0.344,0.356] |
| 4 | 0.082 | [0.079,0.086] | 0.027 | [0.025,0.029] | 0.117 | [0.113,0.121] | 0.043 | [0.040,0.046] | 0.059 | [0.056,0.062] | 0.086 | [0.082,0.090] | 0.010 | [0.008,0.011] | 0.350 | [0.343,0.356] |
| 5 | 0.079 | [0.076,0.082] | 0.029 | [0.026,0.031] | 0.120 | [0.116,0.124] | 0.042 | [0.040,0.045] | 0.058 | [0.055,0.061] | 0.091 | [0.087,0.095] | 0.010 | [0.008,0.011] | 0.346 | [0.340,0.353] |
| 6 | 0.078 | [0.074,0.081] | 0.026 | [0.024,0.028] | 0.116 | [0.112,0.120] | 0.042 | [0.040,0.045] | 0.057 | [0.054,0.060] | 0.090 | [0.086,0.094] | 0.008 | [0.007,0.010] | 0.343 | [0.337,0.349] |
| 7 | 0.078 | [0.075,0.082] | 0.028 | [0.026,0.030] | 0.114 | [0.109,0.118] | 0.042 | [0.040,0.045] | 0.056 | [0.053,0.059] | 0.096 | [0.092,0.099] | 0.010 | [0.008,0.011] | 0.336 | [0.329,0.342] |
| 8 | 0.079 | [0.075,0.082] | 0.025 | [0.023,0.027] | 0.113 | [0.109,0.117] | 0.042 | [0.039,0.045] | 0.060 | [0.056,0.063] | 0.095 | [0.091,0.099] | 0.009 | [0.008,0.011] | 0.333 | [0.327,0.339] |
| 9 | 0.075 | [0.072,0.079] | 0.025 | [0.023,0.027] | 0.112 | [0.108,0.116] | 0.041 | [0.038,0.043] | 0.060 | [0.056,0.063] | 0.094 | [0.090,0.098] | 0.010 | [0.008,0.011] | 0.338 | [0.332,0.344] |
| 10 | 0.073 | [0.070,0.077] | 0.023 | [0.021,0.025] | 0.113 | [0.109,0.117] | 0.040 | [0.038,0.043] | 0.056 | [0.053,0.059] | 0.096 | [0.092,0.100] | 0.010 | [0.009,0.012] | 0.340 | [0.334,0.346] |
| 11 | 0.072 | [0.069,0.076] | 0.022 | [0.020,0.024] | 0.113 | [0.109,0.117] | 0.039 | [0.036,0.042] | 0.062 | [0.059,0.065] | 0.095 | [0.092,0.099] | 0.010 | [0.009,0.012] | 0.331 | [0.325,0.337] |
| 12 | 0.071 | [0.067,0.074] | 0.023 | [0.021,0.025] | 0.113 | [0.108,0.117] | 0.039 | [0.036,0.042] | 0.060 | [0.056,0.063] | 0.095 | [0.091,0.099] | 0.010 | [0.008,0.011] | 0.328 | [0.321,0.334] |
| 13 | 0.070 | [0.066,0.073] | 0.025 | [0.023,0.027] | 0.111 | [0.107,0.115] | 0.037 | [0.034,0.040] | 0.059 | [0.056,0.062] | 0.103 | [0.099,0.107] | 0.010 | [0.009,0.011] | 0.323 | [0.316,0.329] |
| 14 | 0.066 | [0.063,0.069] | 0.021 | [0.019,0.023] | 0.106 | [0.102,0.110] | 0.037 | [0.034,0.039] | 0.061 | [0.058,0.064] | 0.098 | [0.094,0.102] | 0.010 | [0.008,0.011] | 0.327 | [0.321,0.333] |
| 15 | 0.067 | [0.063,0.070] | 0.021 | [0.019,0.023] | 0.103 | [0.099,0.107] | 0.035 | [0.032,0.037] | 0.060 | [0.057,0.063] | 0.101 | [0.097,0.105] | 0.009 | [0.008,0.010] | 0.324 | [0.317,0.330] |
| 16 | 0.063 | [0.060,0.067] | 0.021 | [0.019,0.023] | 0.107 | [0.102,0.111] | 0.034 | [0.032,0.037] | 0.061 | [0.057,0.064] | 0.107 | [0.103,0.111] | 0.009 | [0.008,0.010] | 0.319 | [0.313,0.325] |
| 17 | 0.064 | [0.061,0.068] | 0.019 | [0.017,0.021] | 0.102 | [0.097,0.106] | 0.034 | [0.031,0.036] | 0.059 | [0.056,0.063] | 0.105 | [0.101,0.109] | 0.010 | [0.008,0.011] | 0.312 | [0.306,0.319] |
| 18 | 0.056 | [0.053,0.060] | 0.017 | [0.015,0.019] | 0.098 | [0.094,0.103] | 0.033 | [0.031,0.036] | 0.061 | [0.058,0.064] | 0.107 | [0.103,0.111] | 0.009 | [0.008,0.011] | 0.303 | [0.297,0.309] |
| 19 | 0.057 | [0.053,0.060] | 0.016 | [0.014,0.018] | 0.090 | [0.086,0.095] | 0.028 | [0.026,0.031] | 0.062 | [0.059,0.065] | 0.106 | [0.102,0.110] | 0.009 | [0.008,0.011] | 0.296 | [0.290,0.303] |
| 20th (highest) | 0.044 | [0.041,0.047] | 0.014 | [0.012,0.016] | 0.084 | [0.080,0.089] | 0.028 | [0.025,0.030] | 0.063 | [0.060,0.066] | 0.108 | [0.104,0.112] | 0.008 | [0.007,0.010] | 0.281 | [0.275,0.287] |

**Construction of demographic and socioeconomic variables and sensitivity analyses**

To construct covariates such as parental marital status and childhood residency we used information from several registries from statistics Norway. Marital status is available in registries from 1975 and onwards, and we used information on mother's marital status at child age 16 to approximate the child's environment while growing up. Although these registries have information on marriage and divorce status, a drawback is that they do not have information on cohabitants. This means that there will likely be some misclassification of the non-married group as this group will consist of mothers who are single and divorced, but also mothers who have partnered but not married (cohabitants). To address the magnitude of this misclassification we compared marriage rates at child age 16 to marriage rates at the birth of the child by using data from the Medical Birth registry, see Table A3.

Table A3: comparison of mothers union status from birthyear to age 16

| | Union status by child birth | | Union status by child 16 | |
|---|---|---|---|---|
| Birthyear | Married | Non-married | Married | Non-Married |
| 1967 | 94.6 | 5.04 | 86.35 | 13.65 |
| 1968 | 94.41 | 5.59 | 85.81 | 14.19 |
| 1969 | 93.79 | 6.21 | 85.12 | 14.88 |
| 1970 | 93.02 | 6.98 | 84.55 | 15.45 |
| 1971 | 95.05 | 7.95 | 84.17 | 15.83 |
| 1972 | 91.10 | 8.90 | 83.50 | 16.50 |
| 1973 | 90.44 | 9.56 | 82.65 | 17.35 |
| Total | 92.86 | 7.14 | 84.62 | 15.38 |

*Source:* Medical Birth Registry

A majority, 93 %, were registered as married by birth of the child, however this share had fallen to 85 % when the child was 16. Given a higher share of non-intact families one might wonder if the parental income gradient is primarily found among divorced families. We therefore performed a sensitivity analyses were we examined if the relationship between parental income and later health status similar for a subsample of families for which the mother report on being married at birth and where there is no record of the mother ever being divorced or deceased across child age 0-18.

Further, we speculated if father's income were more strongly associated with worse health compared to mother's income. For example, the economic roles of mother and fathers for these cohorts are somewhat different than the typical dual breadwinner model. For some mothers low income might truly reflect a disadvantage position (i.e., indicate that they are low wage workers or have a marginal attachment to the labor market), however low income could also indicate that they were homemaking wives and thus reflect a "high class" position. Thus, mother's income might be a weaker signal of family resources for these cohorts as they are a mix of both high and low status individuals. The results are found in Figure A5.

Figure A5: Share with any disorder for separate measures of mother and father income rank in childhood (panel A) and share with any disorder separate by mother's marital status (panel B)

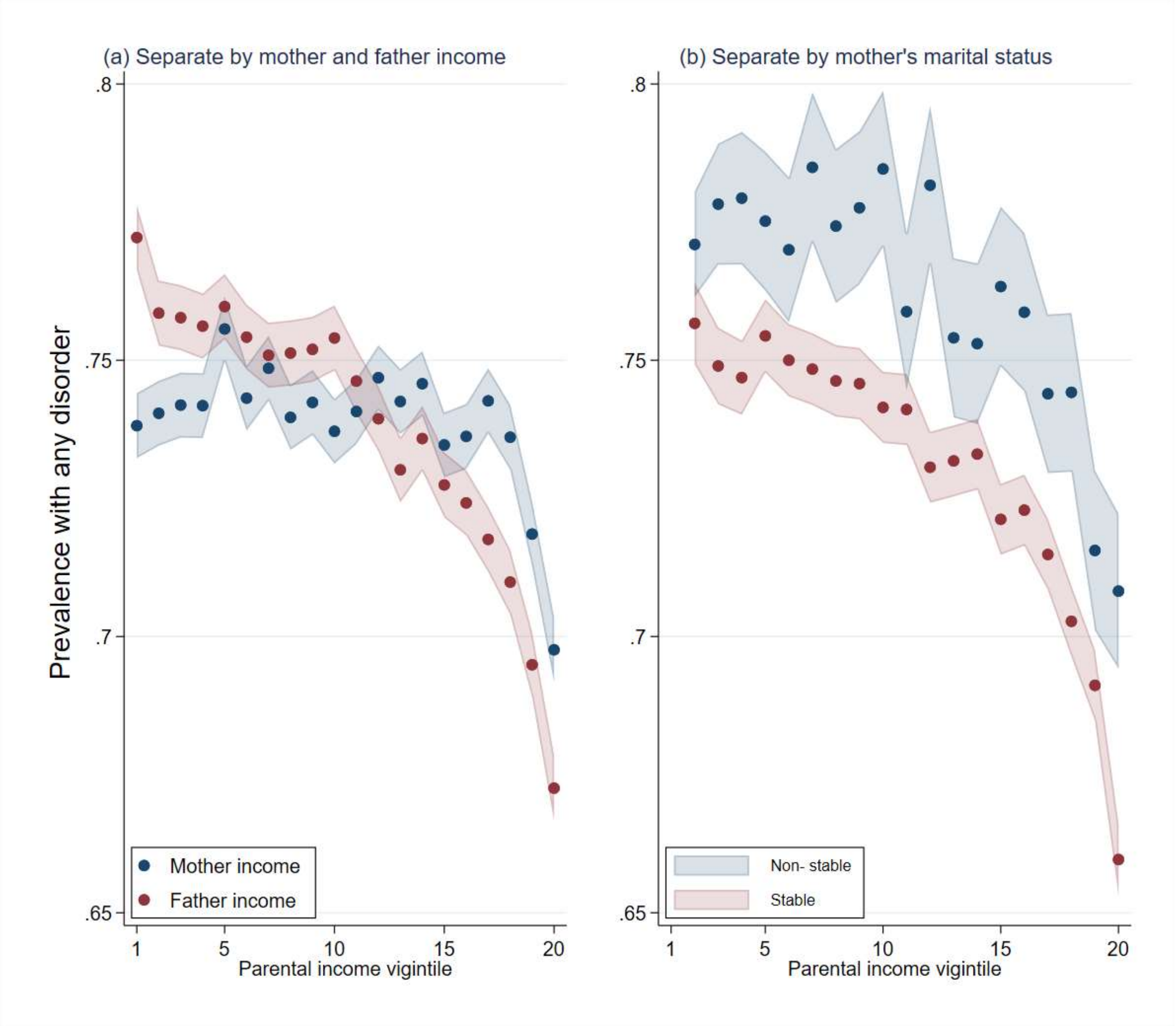


*Source:* Data from Norwegian Control and Reimbursement Database, 2006-2016

We found that father's income, were more strongly associated with worse health at the tails of the distribution, while mothers income were more weakly related to any disorder. Based on marital status we found that, even though individuals of non- married mothers have somewhat higher probability of disorders, the shape is similar for stable married and non-stable married samples, indicating that the gradient in health are not primarily found among non-intact families.

**Robustness of childhood circumstances and parental income gradients in adult health**

We also assessed the extent to which parental income gradients in adult health reflect differences in other childhood circumstances between individuals who grew up in low and high income families. To do so, we examined whether controlling for a set of individuals' childhood factors (i.e., birth weight, parental socio-demographic characteristics, and municipality of residence during adolescence) could account for the observed parental income gradients, similar to an approach used in previous research (2). To achieve this, we first re-estimate equation (1) referred to in the main paper, using a linear specification of parental income (i.e., 0 = bottom percentile, 1 = top percentile) that provides an overall estimate of the income-health association. Then, we estimate this equation while sequentially adding covariates, $\boldsymbol{Z}_i$, for individuals' childhood characteristics:

$$H_i = \alpha_0 + \alpha_1 Parental\ Income_i + \delta \boldsymbol{Z}_i + \mu_i + \varepsilon_i. \quad (1)$$

We then estimate, for the overall health outcome, the percentage change in the coefficient of parental income from adding each set of covariates, $1 - \alpha_1/\beta_1$.

Figure A6 shows results for linear specifications of the parental income gradients measured in percentiles and ranging from 0 to 1 (i.e., 0 = bottom percentile, 1= top percentile) in adult health and how the estimated coefficients decline after adjustments. First, we adjusted for childhood circumstances factors, such as birthweight, geographic region, mother's age at birth and marital status separately, before adjusting for all childhood circumstances combined.

Figure A6: Unadjusted and adjusted association between parental income in percentiles in childhood and adult health at age 39-43 (any disorder), Norwegian birth cohorts 1967-1975

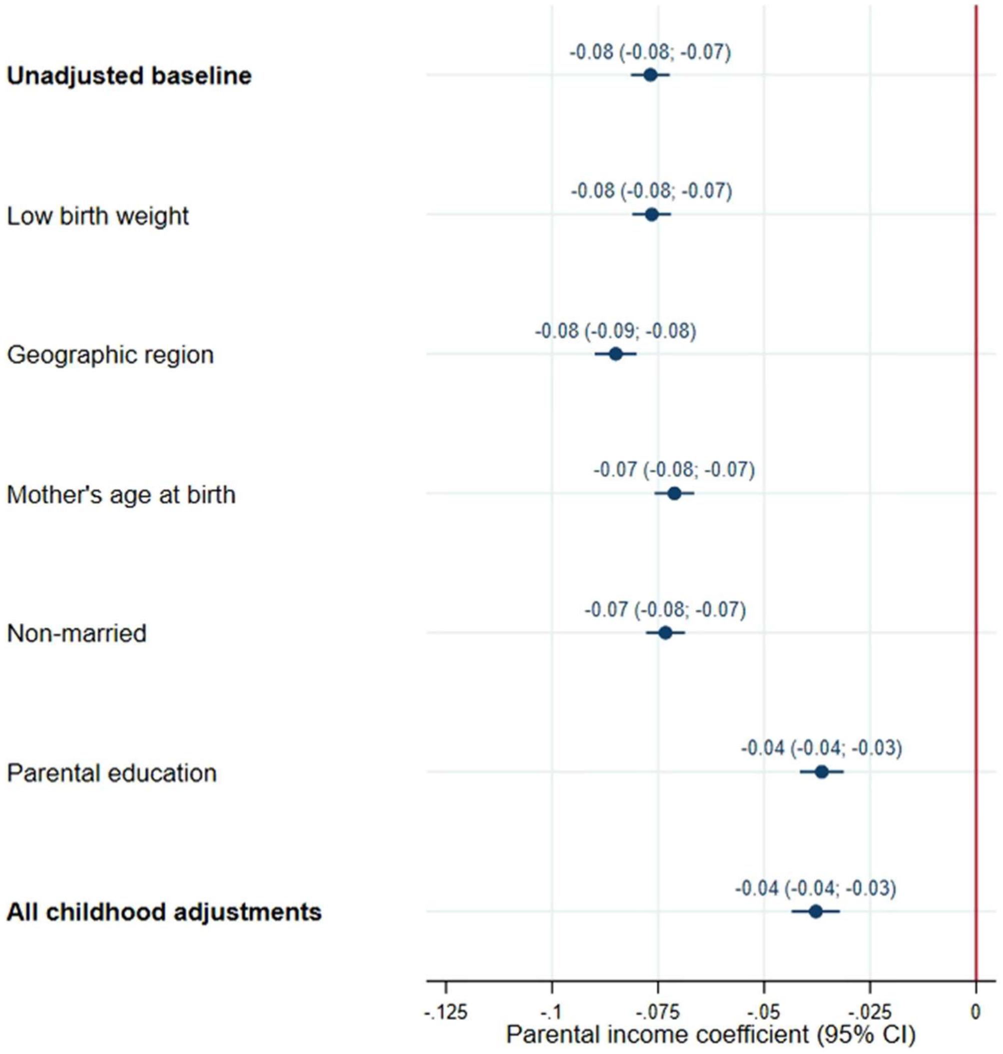


The results show that adjusting for birthweight, mother's marital status, and geographic region (i.e., childhood place of residence) only reduces the parental income coefficient slightly. In contrast, parental education accounts for a larger part of the parental income association.
